# COVID-19 Pandemic among Latinx Farmworker and Non-farmworker Families in North Carolina: Knowledge, Risk Perceptions, and Preventive Behaviors

**DOI:** 10.1101/2020.07.14.20153429

**Authors:** Sara A. Quandt, Natalie J. LaMonto, Dana C. Mora, Jennifer W. Talton, Paul J. Laurienti, Thomas A. Arcury

## Abstract

**Background:** The COVID-19 pandemic poses substantial threats to Latinx farmworkers and other immigrants in food production and processing. Classified as essential, such workers cannot shelter at home. Therefore, knowledge and preventive behaviors are important to reduce COVID-19 spread in the community.

**Methods:** Respondents for 67 families with at least one farmworker (FWF) and 38 comparable families with no farmworkers (non-FWF) in North Carolina completed a telephone survey in May, 2020. The survey queried knowledge of COVID-19, perceptions of its severity, self-efficacy, and preventive behaviors. Detailed data were collected to document household members’ social interaction and use of face coverings.

**Results:** Knowledge of COVID-19 and prevention methods was high in both groups, as was its perceived severity. Non-FWF had higher self-efficacy for preventing infection. Both groups claimed to practice preventive behaviors, though FWF emphasized social avoidance and non-FWF emphasized personal hygiene. Detailed social interactions showed high rates of inter-personal contact at home, at work, and in the community with more mask use in non-FWF than FWF.

**Conclusions:** Despite high levels of knowledge and perceived severity for COVID-19, these immigrant families were engaged in frequent interpersonal contact that could expose community members and themselves to COVID-19.

## 1. Introduction

The 2020 coronavirus pandemic has posed a substantial threat to farmworkers and other workers in the food production and processing system worldwide [1,2]. Such workers are deemed essential workers [3,4] and are unable to practice such preventive measures as sheltering at home that may be recommended to the general population. In addition, food system workers are often of low socioeconomic status, immigrant and minority, undocumented, and living in rural areas so that they are excluded from some of the economic legal protections of workers in other industries [5]. They also may not be reached by public health messaging or provision of personal protective equipment intended to provide them with the knowledge and materials needed to protect themselves [3]. In the United States (US), many immigrant workers lack health insurance and access to health care [6,7], further diminishing their ability to protect their health in a pandemic.

Substantial concern was expressed in the US about Latinx farmworkers’ risk of COVID-19 early in the pandemic [8,9]. These workers often work seasonally, and the Spring work season commenced within the first months of the pandemic. Workers were considered to be at risk because close contact in crowded housing [10,11] and transportation used to reach the fields could increase rates of disease transmission [12,13]. Within the fields, workers often work in close proximity picking row crops; and some equipment requires two or more workers to sit side by side, e.g., on mechanical setters as they plant seedlings. Workers could then act as a vector to their larger communities by infecting other workers and family members. Such patterns were observed in by April, 2020, in immigrant worker populations in meat and poultry processing facilities [14], further increasing the concern for seasonal and migrant crop workers who would begin work in May and June in areas such as North Carolina [15].

Communication about COVID-19 in the US changed over the first few months of the pandemic. Early findings that coronavirus was stable on surfaces for hours or even days [16] led to recommendations that focused on use of cleaning products to sanitize frequently touched surfaces such as door knobs and countertops. These were subsequently downplayed as research and modeling of effects in other countries demonstrated the importance of droplet transmission of the virus, which could be reduced through physical distancing and use of face coverings such as masks [17,18]. Similarly, some early claims for treatment and cures for COVID-19 later proved false or were subject to hurried and incomplete evaluation [19]. Communication of these messages to the public, particularly to those who did not receive communications well in English, sometimes lagged behind scientific findings. Taken together, the changing messages, coupled with public concern, and limited availability of up-to-date information in formats for non-English speakers created a situation in the US in which Latinx workers such as farmworkers were likely to lack accurate information and, as a result, practice ineffective behaviors to protect themselves and prevent spreading disease to their social network.

This study is guided by constructs from the Health Belief Model (HBM) [20]. The HBM tries to understand how knowledge and personal factors lead to actions to protect or promote health. In the HBM, perceptions of one’s susceptibility to a disease and its perceived severity influence actions taken. Individuals must perceive that they are susceptible, in this case, to COVID-19, and that contracting and spreading the disease would have serious consequences. In addition, self-efficacy, the belief in one’s ability to take effective action in the situation of risk to health, influences whether or not one engages in health protective or promoting actions. This suggests that having a strong sense of self-efficacy to practice protective measures to prevent contracting and spreading COVID-19 will lead to engaging in such measures. In this study, we measure a number of these constructs, though we do not execute a full test of the HBM.

We report survey data collected in May 2020 from women in a sample of Latinx farmworker families and a comparison group of Latinx non-farmworker families in North Carolina, USA. The paper has three aims. In all cases, we will compare farmworker families and families with no worker engaged in farm work. First, we will describe the families’ respondents’ (1) knowledge of coronavirus contagion and prevention, (2) risk perceptions, and (3) practices used for prevention and spread of COVID-19. Second, we will describe household social interactions and protections taken, both outside of work and at work. Third, we will use these data to identify specific risks for each group, as well as areas where policy changes can help mitigate the risk for COVID-19.

## 2. Material and Methods

The study reported here is part of a larger two-group, prospective study examining the health and cognitive effects of pesticide exposure in children in farmworker families. The larger study uses a comparative design, with a sample of families of Latinx farmworkers with children and a sample of similar families, but without any farmworker members. Additional details of the study can be found elsewhere [21]. The current study used a telephone survey to reach the mother of the children in these families in May, 2020, when no face-to-face contact between study staff and study participants was permitted by the Institutional Review Board due to COVID-19-related health concerns for research participants. All procedures were approved by the Wake Forest University Institutional Review Board. The study received a Certificate of Confidentiality from the National Institutes of Health.

### 2.1 Inclusion criteria and participant recruitment

Inclusion criteria for the families were similar in both samples when recruited from March, 2018, to December, 2019; they reflect the purpose of the larger study. Each family had to have a child aged 8 years at baseline, who had completed the first grade in the US. All children had to be from families that self-identified as Latino or Hispanic, and with household incomes below 200% of the US federal poverty guideline. In the farmworker sample, the mother or her partner must have been employed in farm work on non-organic farms during the past three years. In the non-farmworker sample, adults could not have been employed in any industry that involves routine exposure to pesticides (e.g., farm work, landscaping, pest control) in the previous three years. Families in the non-farmworker sample could not have lived adjacent to agricultural fields in the previous three years.

Exclusion criteria for both samples included children having life threatening illnesses, prior history of neurological conditions, physical condition or development disorder that would not allow them to complete or would interfere with the results of neurobehavioral tests or MRIs (used in the larger main study), primary language other than Spanish or English spoken in the home, or refusal of mother/guardian to complete the questionnaires.

In the larger study, a total of 76 children were recruited for the farmworker sample and 65 children for the non-farmworker sample. For the recruitment of the original sample, the community partner North Carolina Farmworkers Project developed a list of farmworker families with an 8 year old child, and the locations where they lived. In addition, other community organizations that served farmworker families in the recruitment area were contacted. Study personnel contacted the mothers. Similarly, for the original non-farmworker sample, local recruiters in Winston-Salem, NC, and community members developed a list. For both samples, mothers were contacted by a bilingual staff member who explained the overall study procedures, answered questions, and, if the mother agreed to participate, obtained signed informed consent from the mother and assent from the child. As recruitment progressed, community partners worked with the study team to balance the two samples on socioeconomic status.

Prior to the telephone survey, 5 children in the farmworker sample and 17 in the non-farmworker sample withdrew, moved away from the study area, or were lost to follow-up. The remaining children represented 67 farmworker families and 45 non-farmworker families, because some families had more than one child enrolled. For the telephone sample, 2 families refused to participate and 5 could not be reached, all in the non-farmworker sample. A total of 67 farmworker families and 38 non-farmworker families could be reached and agreed to participate. This sample of 105 is used in this paper.

### 2.2 Data collection

Data for this study were gathered from May 1, 2020, to June 5, 2020, using a telephone survey. Interviewers were members of the larger study team who had usual interview contact with the mothers. Each interviewer participated in an individualized televideo training after which the interviewer was asked to practice completing the form and did an oral practice interview with the study manager. To recruit participants, interviewers called the last known telephone number for the mother in each family, explained the purpose and procedures for the study, and told the mother that she would receive a $10 incentive for completing it at the next in-person study visit. If there was no answer, the interviewers tried at different times of day until the participant was reached or until at least 3 unsuccessful calls had been made.

If the mother agreed to participate, her informed consent was noted, and the interviewer proceeded to conduct a standardized interviewer-administered questionnaire in the language of the participant’s choice using a tablet. Data were entered in real time during the interviews using Research Electronic Data Capture (REDCap). REDCap is hosted at Wake Forest School of Medicine through the Clinical and Translational Science Institute. The REDCap system provides secure, web-based applications for a variety of types of research [22]. Data from this interview were later merged with selected personal, family, and household variables collected in the main study questionnaires.

Questionnaire items relating to the coronavirus and COVID-19 were adapted from existing studies (e.g., McFadden et al. [23]), where available, or from questions recommended for COVID-19 research by governmental and non-governmental agencies. Because of the need for rapid data collection, validation was limited to checks on face validity and interviewer reports of difficulties experienced by respondents during practice interviews.

### 2.3 Variables and measures

Variables from the main study baseline questionnaire were used to create measures to describe the sample. These included the following measures for the mother: age, country of origin, educational attainment, and current occupation. Group assignment of the family to the farm work or non-farm work sample was also noted from the baseline questionnaire. Current household size was obtained by querying the number of adults (persons 18 years and older) and children living in the respondent’s dwelling.

Knowledge of COVID-19 was measured with a series of 4 questions that asked the respondent to identify the correct answer from a series of statements for the definition of COVID-19, its transmission route, the definition of “close contact” for coronavirus, and availability of treatment and vaccine. A summary variable was created by summing the number (0-4) of items answered correctly.

Knowledge of behaviors that can prevent exposure to the coronavirus and its transmission was measured with a set of 13 items in which the respondent was asked whether or not each could prevent exposure for self or others. The list contained 8 items for which the correct response was positive (e.g., wear a face mask when out in public) and 5 items for which the correct response was negative (e.g., take herbal supplements). The number of correct responses was summed to create a summary measure of questions answered correctly, with a range of 0 to 13.

Perceptions of risk was measured with 8 items containing statements about health risk to self and community from COVID-19. Responses used a 5-point Likert-type scale with values ranging from strongly agree to strongly disagree, which was collapsed to a 3-point scale for analysis with values agree, neutral, and disagree. A summary measure of general risk was calculated for the first 6 measures, with disagree/low risk perception given a value of 0, neutral risk perception a value of 1, and agree/higher risk perception a value of 2. These were summed, creating a scale with values of 0 to 12. This was divided into categories of low risk (<8), medium risk (8-9), and high risk (>9). Similarly, two items concerning personal risk or self-efficacy were coded and added to create a scale with values 0 to 4. This was divided into categories of low self-efficacy (0-2) and high self-efficacy (3-4).

Personal behaviors to protect health and prevent spread of the coronavirus in the past month were obtained by asking the respondent if they had never, sometimes, or always practiced each of 10 behaviors. These included the 8 positive behaviors in the knowledge items described above, as well as 2 additional items (avoiding travel to areas infected with coronavirus; avoiding eating outside the home). These were summed with a possible range for the summary being 0 to 20, with each behavior scored as 0 (never), 1 (sometimes), or 2 (always).

To obtain household level measures of social distancing and use of masks for protection, respondents were asked how many adults had visited in the respondent’s house in the past week. Response options were none, 1 or 2, 3 or 4, and 5 or more. Those who had had visiting adults were asked how many visitors had worn masks during their visit, with the response options of all of them, some of them, and none of them. These questions were also asked about child visitors. Respondents were also asked how many different houses, apartments, or trailers of others they had visited in the last week. Response options were none, 1 or 2, 3 or 4, and 5 or more. Those who had visited other homes were asked how often they wore a mask during their visit, with the response options of all, some, or none of the time. Similar questions were asked for children, a spouse/partner, or other adult living in the household. Respondents were asked how many people they worked with, defined as the number of persons with whom they worked closely enough to have a normal conversation for at least some of the work time. Response options were none, 1 or 2, 3 or 4, and 5 or more. Mask use was queried for co-workers, with response options of all of them, some of them, and none of them wore masks at work. Similar questions were asked for the spouse/partner at work. Respondents were asked if their children had been cared for in the past week at a day care, pre-school, school, after school program, or at a relative or friend’s house. Any positive responses were followed by questions of whether all, some or no childcare workers wore masks and wore gloves.

To obtain information on large social gatherings in the past week, respondents were asked if any household member had attended church, the approximate number of attendees, and if all, some or none of the attendees wore masks. The same set of questions was asked about whether any household member had attended a party or other social event such as a cookout, baptism, quinceañera, wedding or funeral in the past week..

### 2.4 Analysis

Frequencies and percents were calculated to examine the variables of interest by farmworker status and significant differences were examined using Chi-Square or Fisher’s Exact tests as appropriate. All analyses were done using SAS v 9.4 (SAS Institute, Cary, NC) and p-values < .05 are considered statistically significant.

## 3.0 Results

### 3.1 Description of the sample

Respondents ranged in age from 25 to 47 years (Table 1). About 80% of both samples were born in Mexico; Spanish was the preferred language for most. Years of formal education for the respondents ranged from 0 to college graduate, with the median in both samples being ninth grade. Their spouse/partners had slightly lower education; the medians for the farmworker and non-farmworker samples were sixth and eighth grades, respectively.

**Table 1.** Individual and household characteristics of participants. Comparison of Latinx farmworker and nonfarmworker adults in North Carolina, May 2020.

|  | Farmworker<br>n=67 |  | Non-Farmworker<sup>1</sup><br>n=38 |  |
| --- | --- | --- | --- | --- |
|  | n | % | n | % |
| Age |  |  |  |  |
| 25 – 29 years | 7 | 10.45 | 5 | 13.16 |
| 30 – 34 years | 26 | 38.80 | 7 | 18.42 |
| 35 – 39 years | 19 | 28.36 | 13 | 34.21 |
| 40 – 47 years | 15 | 22.39 | 13 | 34.21 |
| Country of birth (mother) |  |  |  |  |
| Mexico | 54 | 80.60 | 30 | 78.95 |
| El Salvador | 7 | 10.45 | 0 | 0 |
| Guatemala | 2 | 2.99 | 1 | 2.64 |
| Honduras | 1 | 1.49 | 3 | 7.89 |
| United States | 3 | 4.48 | 2 | 5.26 |
| Other | 0 | 0 | 2 | 5.26 |
| Language most comfortable for conversation |  |  |  |  |
| Spanish | 65 | 97.01 | 35 | 92.11 |
| English | 1 | 1.49 | 3 | 7.89 |
| An indigenous language | 1 | 1.49 | 0 | 0 |
| Highest level of education completed (mother) |  |  |  |  |
| Less than sixth grade | 13 | 19.40 | 3 | 7.89 |
| Sixth - eighth grade | 18 | 26.87 | 8 | 21.05 |
| Ninth – eleventh grade | 25 | 37.31 | 16 | 42.11 |
| High school or more | 11 | 16.42 | 11 | 28.95 |
| Highest level of education completed (spouse)<sup>1</sup> |  |  |  |  |
| Less than sixth grade | 13 | 23.64 | 7 | 19.44 |
| Sixth - eighth grade | 17 | 30.91 | 11 | 30.56 |
| Ninth – eleventh grade | 20 | 36.36 | 7 | 19.44 |
| High school or more | 5 | 9.09 | 11 | 30.56 |

<sup>1</sup> Totals 55 and 36, respectively, due to missing values.

Total household size ranged from 1 to 10 (median=5) and 3 to 13 (median=6) in the farmworker and non-farmworker samples, respectively. For the farmworker sample, the number of adults in the household ranged from 1 to 6, while the number of children ranged from 0 (a respondent currently separated from her family) to 7. For the non-farmworker samples, the ranges were 1 to 4 for adults and 1 to 10 for children.

At baseline, farmworker families reported that the most common industry in which women worked was agriculture; for men, it was construction, followed by agriculture. For non-farmworker families, most women were not in the labor force and the majority of men worked in construction.

### 3.2 Individual Knowledge, Risk Perception and Behaviors of COVID-19

Knowledge of the coronavirus was high (Table 2). All individuals in both samples had heard of the virus, and none required an explanation of what it was. The farmworker sample had more correct answers than the non-farmworker sample on three of the four remaining items. More in the farmworker sample knew that COVID-19 was a respiratory disease caused by a viral infection (100% vs. 89.47%; p<.05). For the item concerning treatment or vaccine for COVID-19, 28.95% of the non-farmworker sample did not know that there is currently no cure or a vaccine for COVID-19, compared to only 5.97% of the farmworker sample (p<.01). Overall, knowledge in the farmworker sample was significantly higher than in the non-farmworker sample (p<.0001), with 94.03% of farmworker sample having a perfect score, compared to only 60.53% of the non-farmworker sample.

**Table 2.** Knowledge of COVID-19. Comparison of Latinx farmworker and nonfarmworker adults in North Carolina, May 2020. Correct responses are italicized.

| Variable | Farmworker<br>n=67 |  | Non-Farmworker<br>n=38 |  |
| --- | --- | --- | --- | --- |
|  | n | % | n | % |
| Are you aware of the coronavirus pandemic? It is sometimes called the COVID-19 pandemic. |  |  |  |  |
| Yes | 67 | 100 | 38 | 100 |
| No | 0 | 0 | 0 | 0 |
| Which of the following three statements is correct about the definition of COVID-19, the disease that results from the coronavirus? <sup>1</sup> |  |  |  |  |
| *Coronavirus is a respiratory disease caused by a viral infection.* | 67 | 100 | 34 | 89.47 |
| The most obvious symptoms usually include respiratory symptoms accompanied by fever, but coronavirus is NOT contagious. | 0 | 0 | 1 | 2.63 |
| Coronavirus can progress to a severe illness, but NEVER leads to death. | 0 | 0 | 3 | 7.89 |
| Which of the following is correct about transmission route of coronavirus? |  |  |  |  |
| *Coronavirus is transmitted through coughing or sneezing.* | 67 | 100 | 37 | 97.37 |
| Coronavirus is NOT transmitted by close contact with people. | 0 | 0 | 1 | 2.63 |
| Which of the following is correct about “close contact” of coronavirus? <sup>1</sup> |  |  |  |  |
| *“Close contact” involves a direct contact with persons’ respiratory secretions.* | 67 | 100 | 35 | 92.11 |
| Relatives and healthcare workers are excluded from the category of close contact. | 0 | 0 | 3 | 7.89 |
| Which one is correct about the treatment or a vaccine for the COVID-19? <sup>2</sup> |  |  |  |  |
| There is a treatment for COVID-19 that cures a patient. | 2 | 2.99 | 2 | 5.26 |
| *Currently, there is neither a cure nor a vaccine.* | 63 | 94.03 | 27 | 71.05 |
| Currently, there isn’t a cure, but there is a vaccine. | 1 | 1.49 | 7 | 18.42 |
| Don’t know | 1 | 1.49 | 2 | 5.26 |

<sup>1</sup>p<.05; <sup>2</sup>p<.01 p-values for the association between farmworker status and correct/incorrect (collapsed)

Knowledge of behaviors to prevent exposure to the coronavirus or spread of COVID-19 was high in both samples (Table 3). For seven of the 13 items, both samples had 100% correct responses. More in the farmworker sample knew that avoiding touching the face with unwashed hands was protective than in the non-farmworker sample (98.51% vs. 84.21%; p<.01). The only other items for which the samples had different responses were three of the five in the list that were negative options (e.g., taking herbal supplements). For these the non-farmworker sample had significant more correct responses for using herbal supplements (55.26% vs. 4.48%; p<.0001). The farmworker sample had more correct responses for eating a balanced diet (68.66% vs 44.74%; p<.05), and getting regular exercise (71.64% vs. 39.47%; p<01). Overall, the farmworker sample had somewhat better knowledge of prevention than did the non-farmworker sample, but the difference was not significant (p=.0562).

**Table 3.** Knowledge of behaviors that can prevent exposure to the coronavirus or contracting COVID-19. Comparison of number and percentage of correct reponses, Latinx farmworker and nonfarmworker adults in North Carolina, May 2020.

| Variable | Farmworker<br>n=67 |  | Non-Farmworker<br>n=38 |  |
| --- | --- | --- | --- | --- |
|  | n | % | n | % |
| Frequent hand washing | 67 | 100 | 38 | 100 |
| Avoid touching your eyes, nose, and mouth with unwashed hands <sup>2</sup> | 66 | 98.51 | 32 | 84.21 |
| Use disinfectants like Clorox or Lysol on frequently touched surfaces like door knobs and counters | 67 | 100 | 38 | 100 |
| Avoid eating meat* | 48 | 71.64 | 33 | 86.84 |
| Stay home when you are sick | 67 | 100 | 38 | 100 |
| Take herbal supplements*<sup>3</sup> | 3 | 4.48 | 21 | 55.26 |
| Cover your cough | 67 | 100 | 38 | 100 |
| Eat a balanced diet*<sup>1</sup> | 46 | 68.66 | 17 | 44.74 |
| Avoid close contact with people who do not live with you | 67 | 100 | 38 | 100 |
| Avoid crowds of people | 67 | 100 | 38 | 100 |
| Get the flu shot* | 39 | 58.21 | 16 | 42.1 |
| Get regular exercise*<sup>2</sup> | 48 | 71.64 | 15 | 39.47 |
| Wear a face mask when out in public | 67 | 100 | 38 | 100 |

\*These responses are NOT effective preventive behaviors, so negative responses were considered correct.
<sup>1</sup>p<.05; <sup>2</sup> p<.01; <sup>3</sup> p<.001

The farmworker sample respondents perceived lower risk associated with COVID-19 for themselves and their community than did the non-farmworker sample respondents (Table 4), with about half of the summed farmworker responses in the middle risk category, and almost two-thirds of the summed responses for the non-farmworkers falling in the highest perceived risk category (p=.0034). Similarly, the farmworker sample perceived that they had lower ability to protect themselves from the coronavirus, with almost all responses (97.01%) falling in the lower self-efficacy category, compared to 73.68% of the non-farmworker sample falling in the higher self-efficacy category (p<0001).

**Table 4.**
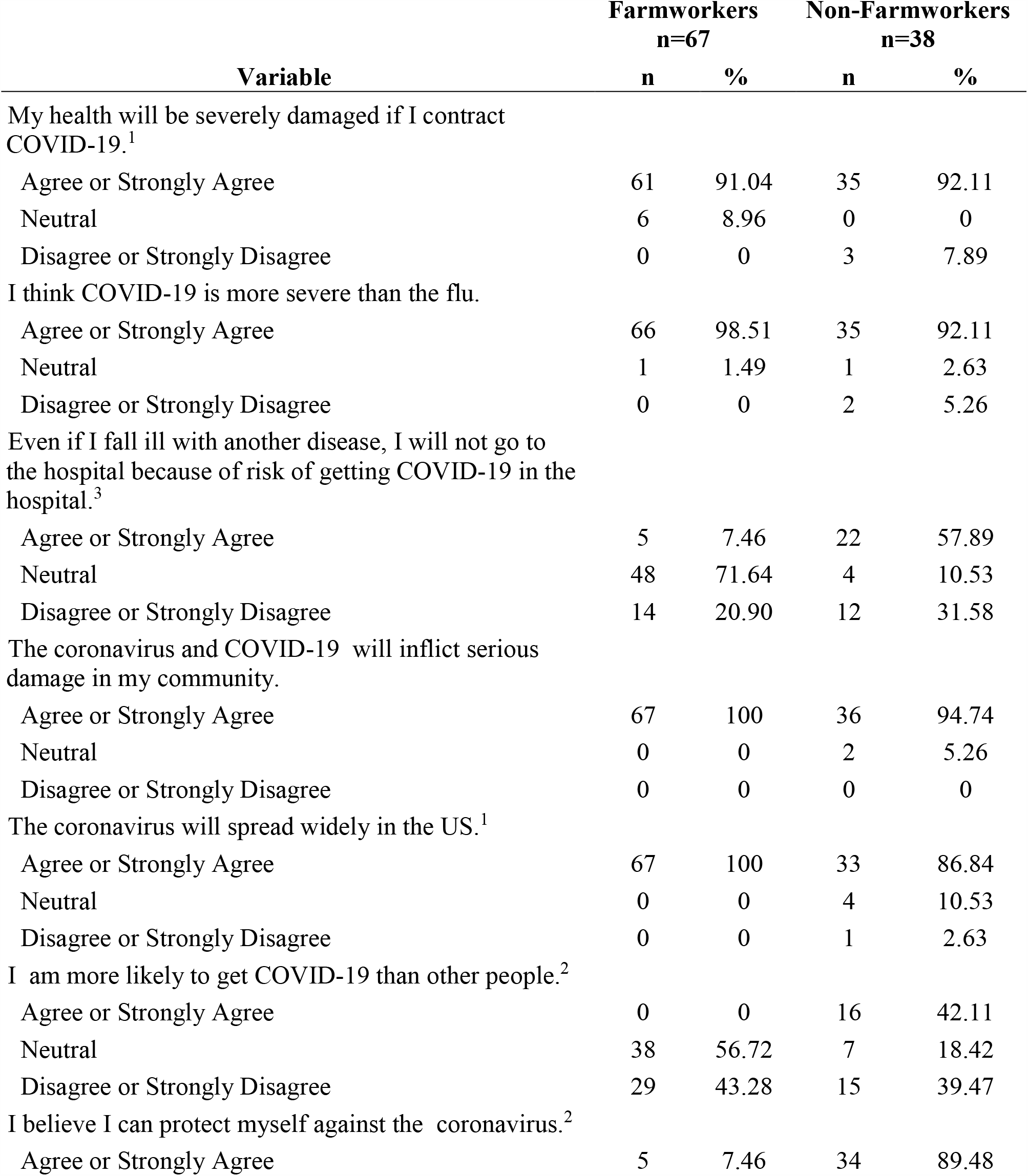
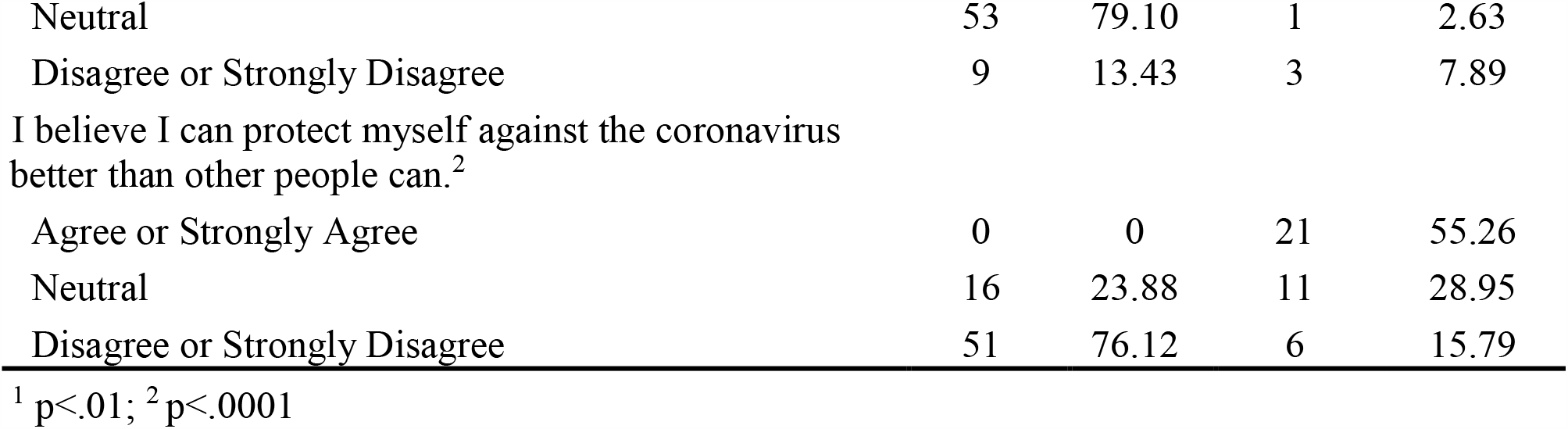
Perceived risks associated with COVID-19. Comparison of percentage of correct reponses, Latinx farmworker and nonfarmworker adults in North Carolina, May 2020.

| Variable | Farmworkers<br>n=67 |  | Non-Farmworkers<br>n=38 |  |
| --- | --- | --- | --- | --- |
|  | n | % | n | % |
| My health will be severely damaged if I contract COVID-19.<sup>1</sup> |  |  |  |  |
| Agree or Strongly Agree | 61 | 91.04 | 35 | 92.11 |
| Neutral | 6 | 8.96 | 0 | 0 |
| Disagree or Strongly Disagree | 0 | 0 | 3 | 7.89 |
| I think COVID-19 is more severe than the flu. |  |  |  |  |
| Agree or Strongly Agree | 66 | 98.51 | 35 | 92.11 |
| Neutral | 1 | 1.49 | 1 | 2.63 |
| Disagree or Strongly Disagree | 0 | 0 | 2 | 5.26 |
| Even if I fall ill with another disease, I will not go to the hospital because of risk of getting COVID-19 in the hospital.<sup>3</sup> |  |  |  |  |
| Agree or Strongly Agree | 5 | 7.46 | 22 | 57.89 |
| Neutral | 48 | 71.64 | 4 | 10.53 |
| Disagree or Strongly Disagree | 14 | 20.90 | 12 | 31.58 |
| The coronavirus and COVID-19 will inflict serious damage in my community. |  |  |  |  |
| Agree or Strongly Agree | 67 | 100 | 36 | 94.74 |
| Neutral | 0 | 0 | 2 | 5.26 |
| Disagree or Strongly Disagree | 0 | 0 | 0 | 0 |
| The coronavirus will spread widely in the US.<sup>1</sup> |  |  |  |  |
| Agree or Strongly Agree | 67 | 100 | 33 | 86.84 |
| Neutral | 0 | 0 | 4 | 10.53 |
| Disagree or Strongly Disagree | 0 | 0 | 1 | 2.63 |
| I am more likely to get COVID-19 than other people.<sup>2</sup> |  |  |  |  |
| Agree or Strongly Agree | 0 | 0 | 16 | 42.11 |
| Neutral | 38 | 56.72 | 7 | 18.42 |
| Disagree or Strongly Disagree | 29 | 43.28 | 15 | 39.47 |
| I believe I can protect myself against the coronavirus.<sup>2</sup> |  |  |  |  |
| Agree or Strongly Agree | 5 | 7.46 | 34 | 89.48 |
| Neutral | 53 | 79.10 | 1 | 2.63 |
| Disagree or Strongly Disagree | 9 | 13.43 | 3 | 7.89 |
| I believe I can protect myself against the coronavirus better than other people can.<sup>2</sup> |  |  |  |  |
| Agree or Strongly Agree | 0 | 0 | 21 | 55.26 |
| Neutral | 16 | 23.88 | 11 | 28.95 |
| Disagree or Strongly Disagree | 51 | 76.12 | 6 | 15.79 |

<sup>1</sup> p<.01; <sup>2</sup> p<.0001

For self-reported actual preventive behaviors, the farmworker sample was significantly more likely to report practicing three behaviors (avoiding travel to areas infected with coronavirus [p<.01], avoiding eating outside the home [p<.01], and avoiding close contact with people who were sick [p<.05]), while the non-farmworker sample was significantly more likely to report four behaviors (washing hands for 20 seconds [p<.001], using surface disinfectants [p<.0001], avoiding touching face with unwashed hands [p<.0001], and covering cough with tissue [<.0001]) (Table 5). The overall difference between the two samples was significant (p=.0008).

**Table 5.** Self-reported frequency of taking measures to prevent infection with the coronavirus in the past month. Comparison of percentage of reponses, Latinx farmworker and nonfarmworker adults in North Carolina, May 2020.

| Variable | Farmworkers<br>n=67 |  | Non-Farmworker<br>n=38 |  |
| --- | --- | --- | --- | --- |
|  | n | % | n | % |
| Avoided travel to areas infected with coronavirus<sup>2</sup> |  |  |  |  |
| Always | 64 | 95.52 | 29 | 76.32 |
| Sometimes | 3 | 4.48 | 4 | 10.53 |
| Never | 0 | 0 | 5 | 13.16 |
| Washed hands with soap and water for 20 seconds<sup>3</sup> |  |  |  |  |
| Always | 39 | 58.21 | 35 | 92.11 |
| Sometimes | 28 | 41.79 | 3 | 7.89 |
| Never | 0 | 0 | 0 | 0 |
| Used disinfectants on frequently touched surfaces<sup>4</sup> |  |  |  |  |
| Always | 13 | 19.40 | 32 | 84.21 |
| Sometimes | 50 | 74.63 | 5 | 13.16 |
| Never | 4 | 5.97 | 1 | 2.63 |
| Avoided touching your eyes, nose, and mouth with unwashed hands<sup>4</sup> |  |  |  |  |
| Always | 15 | 22.39 | 25 | 65.79 |
| Sometimes | 52 | 77.61 | 7 | 18.42 |
| Never | 0 | 0 | 6 | 15.79 |
| Avoided eating outside of the home<sup>2</sup> |  |  |  |  |
| Always | 61 | 91.04 | 27 | 71.05 |
| Sometimes | 6 | 8.96 | 8 | 21.05 |
| Never | 0 | 0 | 3 | 7.89 |
| Stayed home when you were sick. |  |  |  |  |
| Always | 67 | 100 | 36 | 94.74 |
| Sometimes | 0 | 0 | 1 | 2.63 |
| Never | 0 | 0 | 1 | 2.63 |
| Covered your cough or sneeze with a tissue, then threw the tissue in the trash.<sup>4</sup> |  |  |  |  |
| Always | 20 | 29.85 | 29 | 76.32 |
| Sometimes | 45 | 67.16 | 6 | 15.79 |
| Never | 2 | 2.99 | 3 | 7.89 |
| Avoided close contact with people who were sick.<sup>1</sup> |  |  |  |  |
| Always | 66 | 98.51 | 35 | 92.11 |
| Sometimes | 1 | 1.49 | 0 | 0 |
| Never | 0 | 0 | 3 | 7.89 |
| Avoided crowds of people |  |  |  |  |
| Always | 61 | 91.04 | 33 | 86.84 |
| Sometimes | 6 | 8.96 | 5 | 13.16 |
| Never | 0 | 0 | 0 | 0 |
| Wore a face mask when out in public |  |  |  |  |
| Always | 61 | 91.04 | 33 | 86.84 |
| Sometimes | 6 | 8.96 | 5 | 13.16 |
| Never | 0 | 0 | 0 | 0 |

### 3.3 Household Social Interactions and Protections Taken: Outside of Work, at Work, and Social Group Events

Slightly fewer than half of farmworker families (n=31; 46.27%) reported that they had had adult visitors at their home in the past week. Of these, 30 reported that none of the visitors had worn a mask. Similarly, 28 of these families (41.79%) reported that children had visited in their home and none had worn masks. For non-farmworker families, more had had adult visitors (n=21; 55.26%), but some (n=6; 28.57%) had worn masks. A lower proportion of the non-farmworker families had had child visitors (n=14; 36.84%) and some (n=5; 35.71%) had worn masks. More farmworker than non-farmworker family respondents reported visiting the homes of others in the past week (38.81% vs. 23.68%). Both categories of respondents reported visiting 1 or 2 other homes, except 2 from farmworker families who reported visiting 3 or 4. None of the respondents from farmworker families reported wearing masks when visiting; 22.22% of the non-farmworker respondents reported ever wearing masks while visiting.

Respondents from 40.30% of farmworker families reported that their children visited other homes in the past week, and none wore masks. They also reported that 38.98% of their spouse/partners visited other homes and none ever wore masks. Respondents from non-farmworker families reported fewer children (23.68%) and spouse/partners (27.78%) visited other houses, with one spouse/partner visiting 5 or more houses. About a third (30.00%) of spouses were reported to have worn masks, though several respondents did not know, and 66.67% reported their children had never worn masks while visiting other homes.

Similar response patterns were obtained for other adults who lived in the home. Among farmworker families, 23.88% had another adult living in the household, and 43.75% of them had visited other homes, with none wearing masks. Among non-farmworker families, 31.58% had another adult resident in the home; 16.67% had visited other homes, and half had worn a mask.

Among respondents in farmworker families, 31 (46.27%) reported working in the past week. Most (83.87%) worked in places with 5 or more employees in close enough contact to have a normal conversation at least some of the time. These respondents reported that all (86.67%) or some (10.00%) wore masks in the workplace. Almost all of their spouse/partners worked (96.61%); 76.27% worked in places with 5 or more employees in close contact, and some or all wore masks in 60.71%. Similar patterns were reported for other adults living with the farmworker family respondents. About the same proportion of respondents in non-farmworker families worked (44.74%), but fewer (58.82%) worked in places with 5 or more workers in close contact. Most of these respondents reported that all (50%) or some (31.25%) of co-workers wore masks. Almost all (88.89%) spouses worked, though only about half (43.75%) worked in close contact with 5 or more workers. In about three-quarters of these worksites (76.00%), some (12.00%) or all (64%) workers wore masks.

During the time women were surveyed, schools were closed and no children attended preschools or day care centers. Seven (10.45%) of respondents in farmworker families reported that their children were cared for at a friend or relative’s house and that none of the caregivers wore masks or gloves. Four (10.53%) respondents in non-farmworker families reported similar childcare arrangements. However, half reported the caregiver wore masks and gloves.

Five (7.46%) of the respondents in farmworker families reported that a household resident had attended church in the past week. Total church attendance was estimated at 25 (2 cases), 30 (1 case) and 40 (2 cases). All attendees wore masks in 4 of these church services, and none wore masks in the other. Only one respondent among non-farmworker families reported that a household member had attended church in the past week. Attendance was about 10 people and all reportedly wore masks.

Nine (13.43%) respondents in farmworker families reported that a household member had attended a party or social event in the past week. Estimates of total attendees ranged from 10 to 35; none wore masks. By comparison, three (7.89%) respondents in non-farmworker families reported someone had attended a party or social event. In two cases, attendance was estimated at 10; the other was estimated at 20. No one wore masks at 2 of these events.

## 4.0 Discussion

This study was designed to describe the knowledge, perceived risk and susceptibility, and preventive behaviors reported by Latinx immigrant farmworker and non-farmworker families in North Carolina during the first months of the COVID-19 pandemic These families are of particular concern because the rates of COVID-19 nationally are elevated in minority populations. Specifically in North Carolina, on June 1, 2020, Hispanics were reported to make up 10% of the state’s population, but 39% of the state’s COVID-19 cases [24]. At the same time, several farmworker camps were listed as locations of COVID-19 outbreaks by the state Department of Health and Human Services.

The study found that levels of knowledge were high among the Latinx families surveyed, both farmworker and non-farmworker. All respondents had heard of the pandemic and knew what COVID-19 is and how it is transmitted. They had less accurate knowledge about the availability of a cure or vaccine; and women in farmworker families had, overall, slightly more accurate knowledge than did the women in non-farmworker families. Both samples had strong knowledge of the health behaviors that could protect against exposure to the coronavirus and contracting or transmitting COVID-19. In particular, they knew the primary public health messages promoted early in the pandemic. They were less accurate in differentiating these effective behaviors from ineffective behaviors that might be promoted for health risks other than COVID-19, such as exercising and consuming a balanced diet.

Although both groups perceived that COVID-19 presents a serious risk to health, respondents in farmworker families were significantly less likely to affirm personal susceptibility (e.g., that they would avoid going to the hospital for another illness because of risk of contracting COVID-19 there, and that they were more likely than others to get COVID-19). Similarly, these women in farmworker families had lower self-efficacy concerning their ability to protect themselves.

The two samples affirmed different patterns of health promoting behaviors. For the farmworker families, behaviors that entailed avoiding others (e.g., not traveling to areas infected with coronavirus, avoiding eat out, and avoiding close contact with sick individuals) were affirmed significantly more often than by the non-farmworker families. The latter were more likely to affirm behaviors related to personal hygiene: hand washing, use of disinfectants, avoiding touching the face, and covering coughs and sneezes.

Together, these findings give a sense that, while the women in farmworker families had somewhat better knowledge, they perceived less personal susceptibility to COVID-19. They had low confidence that they could protect themselves. This may be underlying the protective behaviors they reported. They avoided people and places that might be contaminated, but did not subscribe to practicing personal hygiene behaviors. Women in non-farmworker families had greater confidence that they could protect themselves and they claimed to practice more personal hygiene behaviors.

Social desirability [25] can bias the way individuals respond to lists of health behaviors. With knowledge of recommendations, they may tend to see themselves or want to portray themselves as more positive and compliant than they actually are. In order to investigate behaviors in detail and try to avoid social desirability bias, the telephone survey included a series of questions about social interactions by household members and wearing masks. Complex question sequences are thought to reduce social desirability bias [26,27]. The focus on distancing and masks was considered important in light of the developing public health messages that identified the greater importance of maintaining physical distancing and protection against spreading infected droplets with masks, rather than practices such as disinfecting surfaces that had been promoted over mask use earlier in the pandemic [17].

The responses to these questions contrasted sharply with the other reported protective behaviors. They showed a high level of social interaction beyond the immediate household for both farmworker and non-farmworkers families, with both adults and children coming into the homes of respondents and members of the respondent’s household visiting in the homes of others. There was virtually no mask wearing reported by farmworker family respondents, and only some use of masks reported by non-farmworker respondents. Household sizes reported in this study (median 5 for farmworker and 6 for non-farmworker families) are considerably larger than the US average of 2.6 people reported for 2018 [28], making for large potential social networks of contacts.

Many of the adult household members were reported to be working outside the home and working in situations where they had close social contact with other workers. These situations, plus the sheer number of adults in the household (up to 6 in farmworker families and 4 in non-farmworker families), make the potential quite high for additional contacts that could spread the coronavirus. Mask use was reported to be common in the workplaces, though measures of the consistency or enforcement of mask use were not obtained.

The respondents and their family members report continuing to engage in social situations with large numbers in attendance, particularly among the farmworker families. Although masks appear to have been worn for church attendance, little mask wearing was reported for other types of social events.

In total, these results indicate that, despite relatively high knowledge, strong perceptions of risk from COVID-19, and claims of avoiding situations where contracting or spreading infection might be likely, many of the farmworker families included here do not practice safe physical distancing measures as recommended, and use of masks appears to be confined to work settings. The situation for the non-farmworker families appears to be somewhat better, with somewhat greater mask wearing reported, particularly in large social gatherings.

The inconsistency between women in farmworker families seeing themselves as avoiding situations for infection and their actual practices may be due to their living situations and to cultural values. Most live in rural environments and few women drive [29], so that they may perceive of themselves and their households as isolated from population centers. Nonetheless, it is clear that interactions take place within and between households, which can exponentially raise the possibility of transmitting infection. This is in contrast to the non-farmworker families who live in urban environments, many in multi-unit dwellings such as apartment buildings.

They may correctly perceive less ability to socially isolate themselves so give greater importance to personal hygiene measures to prevent infection.

For these immigrant workers, both living in close proximity to extended family members and the cultural value of *familismo* [30] likely affect interpretation of public health recommendations to maintain physical distance. Many immigrant workers settle in the US with extended family from their home communities—siblings, cousins, parents, aunts, and uncles. This can provide considerable social and material support while living in a new environment and working in low wage jobs; family and household boundaries are likely more fluid than they are for other ethnic groups. These relationships are supported and reinforced by *familismo*. This cultural construct includes strong identification with and loyalty to family, as well as respect for family members and placing family needs over one’s own needs. Time spent with one’s immediate and extended family is valued. In such a context, wearing masks or refusing social interaction might be considered an affront. The result can be greater contacts and less physical distancing than public health recommendations intend, increasing the risk of coronavirus infection.

The farmworker families included in this study are seasonal workers, meaning that they live in the area year round and family members work seasonally in agriculture. They may not experience the extremely crowded barracks-style sleeping quarters, kitchens, and bathroom facilities of much of the grower-provided housing where migrant workers live [12]. However, these seasonal worker families do have crowded housing [10,11], and they face worksite hazards for infection in crowded transportation to the fields and working in close quarters in some situations in the fields, as well as in greenhouses or packing facilities. They also often work alongside migrant workers who live in crowded conditions. Although the respondents indicate mask usage, it is difficult to know how sustained that can be, considering the high levels of heat and humidity these workers endure in the fields [31].

This study did not collect data on information sources about COVID-19 available to study participants. Although both groups frequently get information from Spanish language radio, the non-farmworker families may have had greater access to public health signage and other local messages in an urban context than the farmworker families did in rural settings.

Other study limitations include the fact that behaviors were self-reported and not observed. Responses could not be anonymous because they were collected by interviewers that the women had known through participation in the larger study; this could have increased the social desirability in responses concerning behavior. Small sample sizes prevent more detailed analyses of data.

Nevertheless, this study represents a unique opportunity to document the knowledge, perceptions, and behaviors of Latinx immigrants in the US during the early days of the COVID-19 pandemic. In particular, farmworkers are often a hidden and difficult to reach population. This study demonstrates that even with a strong knowledge base, these farmworker families lack the self-efficacy to avoid the coronavirus and COVID-19. While they appear to believe that they are following public health recommendations on physical distancing and wearing masks, detailed data on their social interactions and use of personal protective equipment show that this is not the case. A comparison group of urban-dwelling Latinx immigrants had greater self-efficacy, which might led to the greater use of masks as personal protection reported by respondents in these non-farmworker families.

## 5.0 Conclusions

The transmission of a highly infectious virus like the coronavirus is facilitated by close contact among individuals in a population. The large household sizes, particularly large number of adults working in industries deemed essential, and weak adherence to personal protective equipment such as masks makes the immigrant Latinx population at risk for high rates of infection. It is likely that simple public health messages encouraging physical distancing and mask wearing may not protect the population in the context of structural barriers such as crowded housing and work in essential industries, coupled with strong cultural values placed on support of large extended families. Specific actions beyond what is currently being taken by public health authorities may help improve the health-related behavior reported here and curb the spread of infection in this population. Developing and disseminating culturally sensitive education to help families understand the extent of their social contact and the dangers it poses is essential. Using adult educational approaches that could include interactive exercises to demonstrate the potential spread of infection would likely be more effective than print materials.

The COVID-19 pandemic has ravaged urban populations around the world. While one might expect urban and rural conditions in the US to be markedly different, the findings here suggest that this may not be the case for Latinx workers in essential rural industries. Living in large households and working in close contact with large groups of workers may negate the expected isolation of rural communities.

## Data Availability

Data are available on request.

## Author Contributions

SAQ participated in all aspects of the study conceptualization and design, analysis, and manuscript writing. NJL participated in background research, data coding, and manuscript writing. DCM participated in the participant recruitment, data collection, data management, and manuscript writing. JWT participated in the data analysis, and manuscript writing. PJL participated in all aspects of the study conceptualization and design, and manuscript writing. TAA participated in all aspects of the study conceptualization and design, analysis, and manuscript writing. All authors have read and agreed to the published version of the manuscript.

## Acknowledgments

The authors appreciate the support and participation of their community partner, the NC Farmworkers Project, and of Student Action with Farmworkers. They also appreciate the valuable contributions of our community field interviewers in carrying out participant recruitment and data collection. They especially thank the mothers who participated in this study.

This research was funded by a grant from the National Institute of Environmental Health Science, Grant Number R01 ES08739.

